# H-Type Hypertension and Aneurysm Instability: An Observational and Genetic Study

**DOI:** 10.64898/2026.03.25.26349344

**Authors:** Ji-Wan Huang, Hao Yuan, Rui-Yan Liu, Xue-Yan Deng, Cai-Hong Li, Yu-Xin Li, Bo-Han Cai, Le-Le Dai, Wen-Xin Chen, Chi Huang, Meng-Shi Huang, Zhuo-Hua Wen, Run-Ze Ge, Can Li, Jian-Cheng Lin, Xi-Ru Zhang, Shu-Yin Liang, Can-Zhao Liu, Yi Tu, Yi-Ming Bi, Fa Jin, Shi-Xing Su, Xin Zhang, Xi-Feng Li, Zhi-Bo Wen, Chuan-Zhi Duan, Xin Feng

**Affiliations:** Neurosurgery Center, Department of Cerebrovascular Surgery, Engineering Technology Research Center of Education Ministry of China on Diagnosis and Treatment of Cerebrovascular Disease, Zhujiang Hospital, Southern Medical University, Guangzhou, China; Department of Epidemiology, School of Public Health, Southern Medical University, Guangzhou 510515, Guangdong, China; The Second Clinical Medical School, Southern Medical University, Guangzhou, China; Department of Radiology, Zhujiang Hospital of Southern Medical University, Guangzhou, Guangdong, China; Engineering Research Center of Diagnostic and Therapeutic Technology and Devices for Cerebrovascular Diseases, Ministry of Education; Department of Cardiology, Laboratory of Heart Center, Translational Medicine Research Center, Zhujiang Hospital, Southern Medical University, 253 Gongye Middle Avenue, Guangzhou, Guangdong, 510282, China

**Keywords:** H-type hypertension, intracranial aneurysm, homocysteine, aneurysmal-wall enhancement, Mendelian randomization, prospective cohort study

## Abstract

**Background:** Unruptured intracranial aneurysms (UIAs) pose a significant risk of subarachnoid hemorrhage. Both hypertension and hyperhomocysteinemia are recognized as independent risk factors for vascular disease; however, their combined impact (H-type hypertension) on aneurysm instability and rupture remains unclear.

**Methods:** We analyzed a prospective cohort of 358 adults with UIAs (475 aneurysms) using high-resolution vessel-wall MRI (HRVWI) for cross-sectional and longitudinal assessment. H-type hypertension was defined as hypertension with plasma homocysteine ≥10 μmol/L. Multivariable logistic regression assessed associations with AWE and aneurysm growth (longitudinal sub-cohort: n = 82, 89 aneurysms). Mendelian randomization (MR) analyses evaluated the causal role of homocysteine in hypertension and aSAH. Proteomic profiling identified potential molecular mechanisms.

**Results:** AWE occurred in 33.7% of aneurysms, which were larger, irregular, and had higher PHASES scores. Elevated homocysteine (10.3 vs 9.5 μmol/L, p = 0.004) and H-type hypertension (43.8% vs 28.3%, p < 0.001) were associated with AWE. After adjustment, H-type hypertension independently predicted AWE (OR = 3.18) and aneurysm growth (OR = 3.63). MR analyses showed homocysteine increased aSAH (OR = 1.39) and hypertension risk (OR = 1.10), while hypertension increased aSAH risk (OR = 1.58). Mediation analysis did not support hypertension as a mediator (p = 0.20). Proteomic analyses identified key pathways related to inflammation–immune dysregulation, extracellular matrix remodeling, and signaling activation as potential mediators.

**Conclusions:** H-type hypertension amplifies aneurysmal-wall instability and growth. Combined control of blood pressure and homocysteine merits prospective evaluation for UIA prevention.

## Introduction

Unruptured intracranial aneurysms (UIAs) pose a significant neurovascular risk, with potential rupture leading to severe subarachnoid hemorrhage.^1,2^ Hypertension is a well-documented factor in both aneurysm formation and rupture.^3–5^ Additionally, emerging evidence underscores the role of homocysteine (Hcy) in vascular wall pathology.^6–8^ Both hypertension and hyperhomocysteinemia are recognized as independent risk factors for cardio-cerebrovascular diseases, with their combination may exacerbate vascular damage, leading to an increased risk of aneurysmal rupture.^9–12^

H-type hypertension, characterized by concurrent hypertension and elevated plasma homocysteine levels (≥10 µmol/L), has attracted research attention.^9^ However, its impact on intracranial aneurysm instability and rupture has not been thoroughly investigated. High-resolution magnetic resonance vessel wall imaging (HRVWI) reveals aneurysmal wall enhancement (AWE), a potential marker of aneurysm wall inflammation and instability, linked to increased rupture risk.^13–17^ Previous studies have demonstrated a correlation between elevated serum homocysteine levels and aneurysmal wall enhancement in fusiform aneurysms, suggesting a potential role for homocysteine in aneurysmal pathology.^18^ Nonetheless, the interplay between hypertension and hyperhomocysteinemia, especially their combined influence on aneurysm wall enhancement and the subsequent risk of aneurysm growth and rupture, remains insufficiently elucidated. Mendelian randomization (MR), using genetic variants as instrumental variables, offers a method to explore the causal relationship between homocysteine levels and aneurysm pathology, addressing confounding and reverse causation often found in observational studies.^19^

In this study, we examine the association between H-type hypertension and intracranial aneurysm instability using a dual-method framework that integrates cross-sectional and longitudinal data, along with HRVWI and MR analysis. While the observational component provides estimates of clinical associations, the MR approach offers genetic insights into causality. This complementary strategy strengthens analytical validity, facilitates cross-validation of findings, and helps address the limitations inherent in either method alone.

## Methods

### Observational Analysis

#### Participants and Study Design

This prospective cohort study, registered at ClinicalTrials.gov (NCT06447714), was designed to include both cross-sectional and longitudinal analyses. We consecutively enrolled adults (age ≥18 years) diagnosed with intracranial aneurysms (IAs) who underwent high-resolution vessel wall imaging (HR-VWI) between October 2022 and October 2024. All participants provided written informed consent, and the study protocol was approved by the Institutional Review Board (IRB Number: 2023-KY-038-03). At baseline, demographic characteristics, medical history, and behavioral vascular risk factors were collected using a standardized questionnaire (**detailed in Supplemental Data 1**). A total of 401 patients with 525 IAs were initially screened with HR-VWI. After applying the exclusion criteria, 358 patients with 475 IAs formed the cohort for the cross-sectional analysis of baseline imaging and clinical characteristics. From this cross-sectional cohort, patients who were managed conservatively and had at least one follow-up imaging assessment were selected for longitudinal observation. Consequently, 82 patients with 89 aneurysms were included in the longitudinal analysis to evaluate aneurysm growth. The detailed selection criteria, follow-up protocols, and conservative management rationale for this longitudinal sub-cohort are provided in the **Supplemental Data 2**.

### Inclusion and Exclusion Criteria

Inclusion Criteria: Adults aged 18 years or older diagnosed with IAs who underwent HRVWI. Exclusion Criteria: (1) Previously treated aneurysms; (2) Patients with intracranial aneurysms complicated by other intracranial vascular diseases, such as arteriovenous fistulas, arteriovenous malformations, and Moyamoya disease; (3) Patients with autoimmune diseases, hematological disease, or a history of cancer; (4) Patients with inadequate venous blood samples collected upon admission, poor image quality, or incomplete medical records.

### Data Collection

Demographic characteristics (age, sex, and body mass index [BMI]), medical history (diabetes, hyperlipidemia, aneurysmal subarachnoid hemorrhage [aSAH], ischemic events, and family history of aneurysm), and behavioral vascular risk factors (smoking, alcohol use, lipid-lowering medication and aspirin intake [daily intake of ≥81 mg in the last 3 months]) were collected prospectively via a standardized questionnaire within 12 hours of hospital admission. Aneurysmal characteristics including coexisting with two or multiple aneurysms, location, size, and irregular morphology, and symptomatic aneurysms were recorded. Aneurysms with blebs, daughter sacs, or other types of wall protrusions on 3D images were considered irregular shapes.

Seated resting blood pressure was measured in triplicate using a calibrated electronic sphygmomanometer after a 5-minute rest; the average value was used for analysis. Hypertension was defined as systolic BP ≥140 mmHg and/or diastolic BP ≥90 mmHg, or current use of antihypertensive medication. The use of antihypertensive drugs was recorded. Blood samples were drawn after overnight fasting for the measurement of serum Hcy, and glomerular filtration rate (GFR). Renal function was assessed by the estimated glomerular filtration rate (eGFR) and classified according to the Kidney Disease: Improving Global Outcomes (KDIGO) guidelines as: G1 (≥90 mL/min/1.73 m²), G2 (60-89), G3a (45-59), G3b (30-44), G4 (15-29), and G5 (<15).^20^ serum homocysteine was measured using an enzymatic cycling assay (based on S-adenosylhomocysteine hydrolase) on an automated clinical chemistry analyzer. The assay demonstrates good precision, with intra- and inter-assay coefficients of variation <5%. The cutoff of ≥10 μmol/L was selected to define hyperhomocysteinemia, as it is widely adopted in cardiovascular risk stratification in Asian populations and aligns with regional guideline considerations.^9^ Sensitivity analyses using an alternative cutoff (≥15 μmol/L) were performed to assess the robustness of the findings. H-type hypertension was defined as the presence of both hypertension and Hcy ≥ 10 μmol/L.^9^

### Imaging Measurements

All patients underwent scanning using a 3T MR scanner (Ingenia, Philips, Best, Netherlands) with a 32-channel head coil. The scanning parameters are summarized in **supplemental data 3**. As outlined in our previous studies, the contrast ratio, CRstalk = SIwall / SIstalk, was calculated; AWE was defined as CRstalk ≥ 0.60.^21^ Aneurysms were classified as enhanced or unenhanced based on AWE. A schematic illustration is in **Figure 2**. To standardize AWE assessment and ensure inter-rater reliability, two board-certified neuroradiologists with over 15 years of neurovascular imaging experience each underwent a structured training and calibration process before image evaluation. This process involved a review of established AWE criteria and reference cases, followed by independent assessment of a training image set. Inter-rater agreement was assessed using Cohen’s kappa statistic. During formal evaluation, both readers were blinded to all clinical and biomarker data and independently assessed the presence, pattern, and morphology of AWE and aneurysms. Discrepancies were resolved through consensus; in cases where consensus could not be reached, a third, more senior radiologist made the final determination.

**Figure 1.**
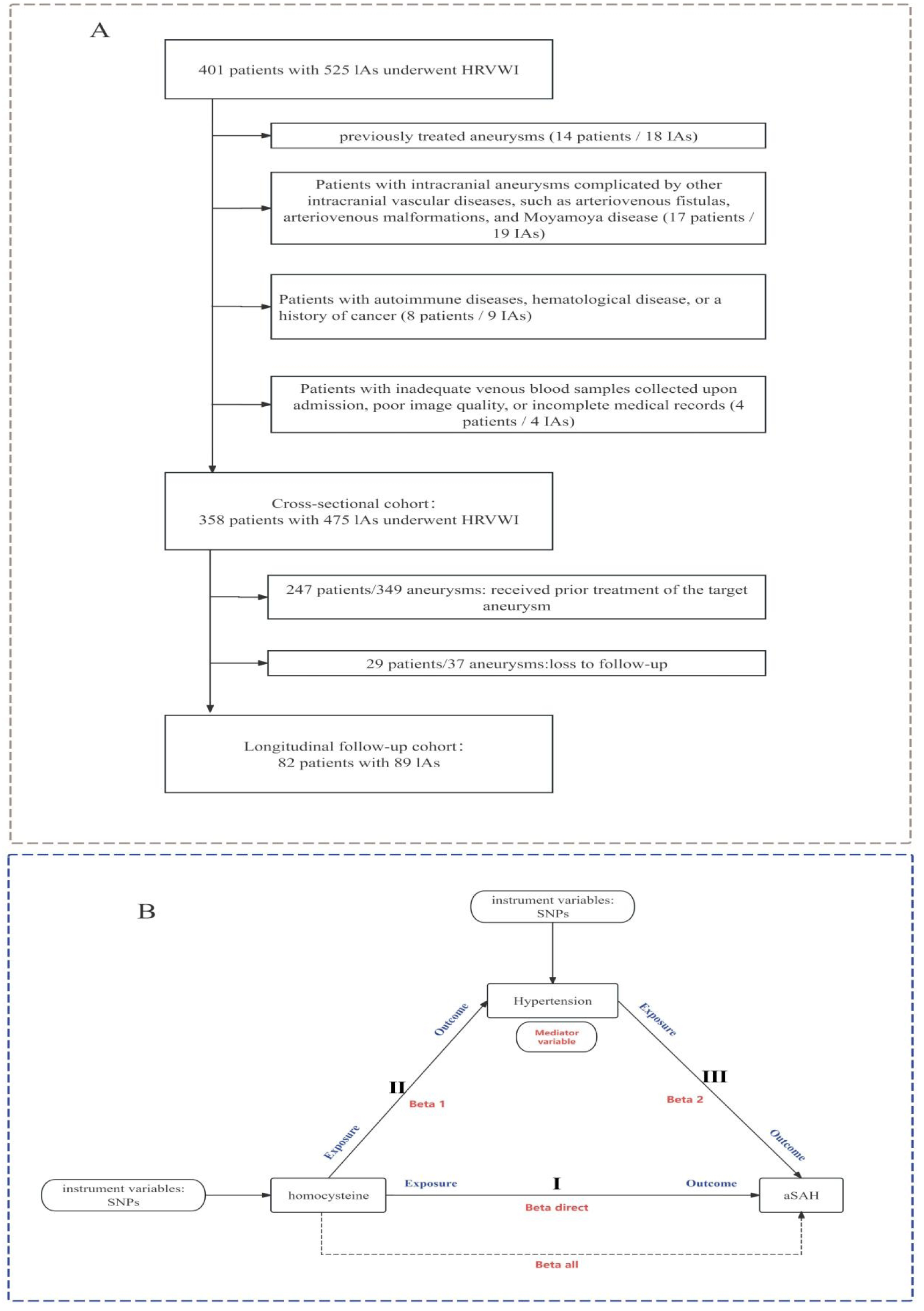
Flow chart showing the general methodological flow of this study. A. Flow diagram of sample selection for cross-sectional cohort study and longitudinal cohort study; B. Mendelian randomization model examining the relationships among homocysteine, hypertension, and aneurysmal subarachnoid hemorrhage (aSAH). Genetic variants (SNPs) influence homocysteine levels, which in turn affect hypertension (pathway II). Both homocysteine and hypertension independently or synergistically contribute to the risk of aSAH (paths I and III). Arrows represent the hypothesized causal pathways, with exposures and outcomes clearly indicated. Beta all, effects of Hcy on aSAH; Beta 1, effects of Hcy on hypertension; Beta2, effects of hypertension on aSAH; Beta direct = Beta all - (Beta 1 * Beta 2). Abbreviation: IAs = intracranial aneurysms; HRVWI = High-resolution magnetic resonance vessel wall imaging; SNP=single-nucleotide polymorphism.

**Figure 2.**
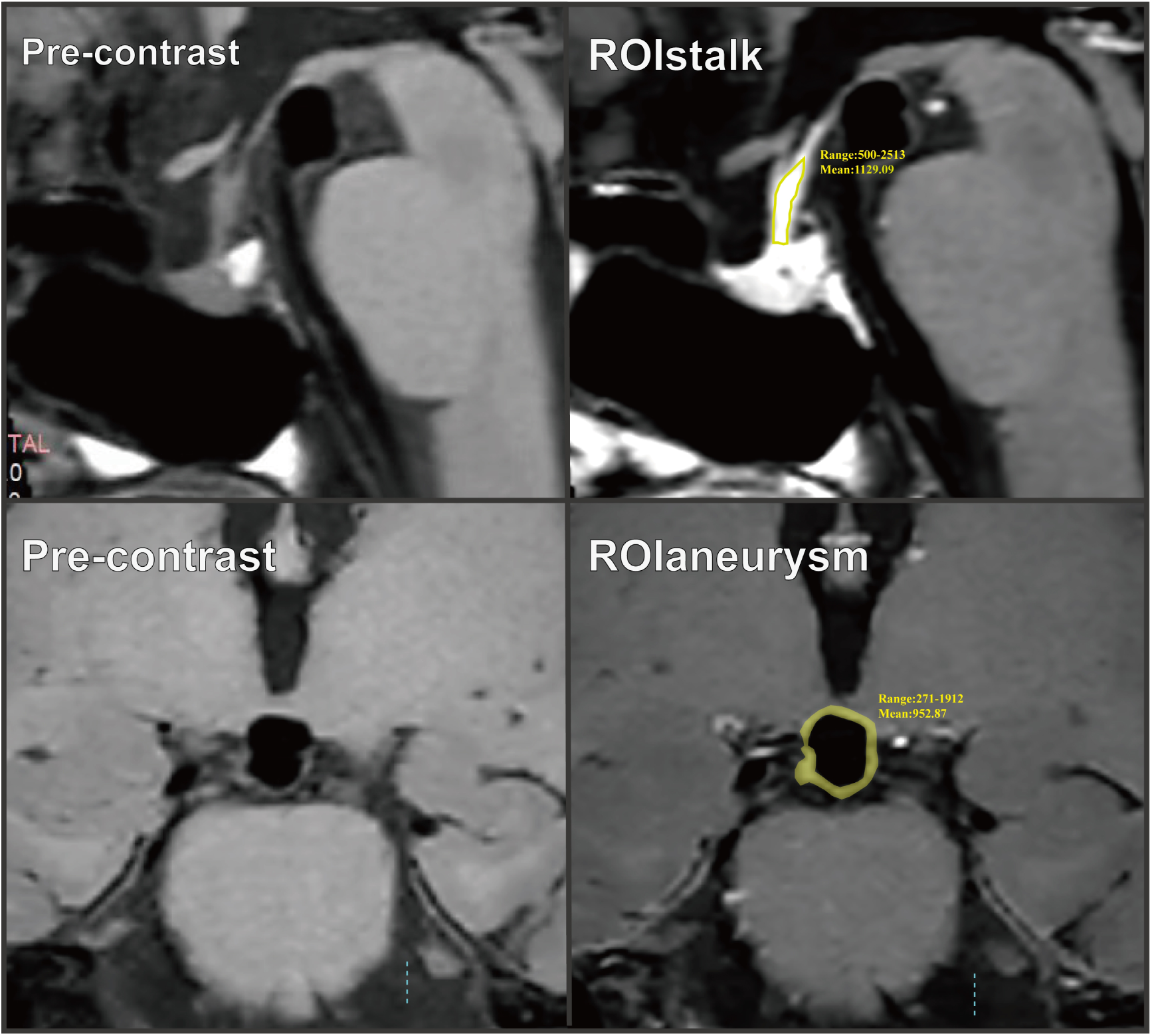
Example case for image measurement. First line, the original image. Second line, the region of interest (ROI) of aneurysm wall and pituitary stalk. The boundaries of the aneurysm wall and the pituitary stalk were outlined manually in the post-contrast T1-weighted images.

### Two-Sample Mendelian Randomization Analysis

We conducted a two-sample Mendelian randomization (TSMR) study to examine the causal relationship between genetically predicted changes in plasma total homocysteine (Hcy) levels and the risk of aneurysmal subarachnoid hemorrhage (aSAH). The validity of the instrumental variables (IVs) in this analysis, represented by the single-nucleotide polymorphisms (SNPs), relies on three key assumptions: (1) a robust association between the SNPs and the exposure; (2) independence of the SNPs from potential confounders affecting the exposure-outcome relationship; and (3) the SNPs influence the outcome solely through the exposure.^22^ Furthermore, given the significant confounding effect of hypertension identified in our clinical observational study on IA wall enhancement, we designated hypertension as a potential mediator. Within this analytical framework, we employed a two-step MR approach to quantify the potential mediating role of hypertension in the causal pathway between Hcy and aSAH.^23^ Since this analysis used summary statistics from publicly available data, no additional ethical approval was required. Details of MR analysis data sources are described in the **supplemental data 4**.

### Proteomics data synopsis

To elucidate the molecular pathways linking homocysteine (Hcy) to aneurysmal subarachnoid hemorrhage (aSAH) and identify potential therapeutic targets, we performed proteomic analyses using a stringent analytical framework consistent with that applied in SNP selection. A two-step Mendelian randomization approach was implemented to evaluate the mediating effects of circulating proteins. Primary pQTL data were derived from the deCODE Health Study, which profiled 4,907 plasma proteins in 35,559 Icelandic participants using the SomaScan platform.^24^ Replication analyses were conducted using two independent cohorts: the UK Biobank Pharma Proteomics Project (UKB-PPP), which measured 2,940 proteins in 54,219 individuals using the Olink platform,^25^ and a Finnish cohort that profiled 3,892 proteins in 10,708 participants using SomaScan.^26^ Cross-cohort and cross-platform validation was performed to ensure the robustness and reproducibility of the identified protein mediators.

### Statistical Analysis

Statistical analyses were performed using R software (version 4.2.2). The normality of continuous variables was assessed using the Kolmogorov-Smirnov test. Normally distributed variables are presented as mean ± SD, while non-normally distributed variables are reported as median [IQR]. Categorical variables are expressed as counts and percentages. Group comparisons for continuous variables were made using the Student’s t-test or Mann-Whitney U test, and categorical variables were compared using the chi-square (χ²) test or Fisher’s exact test.

For cross-sectional analysis, multivariable logistic regression was used to assess the single and joint associations of hyperhomocysteinemia and hypertension with aneurysm wall enhancement (AWE), adjusting for age, sex, BMI, smoking status, aneurysm characteristics (location, size, shape, and presence of multiple aneurysms), eGFR Grade, Lipid-lowering Therapy. Three models were used: Model 1 (unadjusted), Model 2 (adjusted for age, sex, and BMI), and Model 3 (fully adjusted). For the longitudinal analysis on aneurysm growth, the same three models were applied. To address sparse data and potential separation due to small sample size, Firth’s logistic regression was used. Simplified H-type hypertension was converted into a binary variable (presence vs. absence), with adjustmentfor age, sex, BMI, smoking status, aneurysm size, and shape. Results are reported as odds ratios (ORs) with 95% CIs, and statistical significance was set at P < 0.05. Subgroup analyses were conducted based on age (≥70 years), sex, smoking status, alcohol consumption, diabetes, hyperlipidemia, multiple aneurysms, aneurysm size (≥7mm), and location. Sensitivity analyses evaluated the robustness of findings by changing the AWE threshold (CR_stalk ≥0.7), defining hyperhomocysteinemia as Hcy≥15 µmol/L, and excluding participants with a GFR < 60 mL/min/1.73 m².

Mendelian randomization (MR) analyses were conducted using the "TwoSampleMR" package in R (version 4.4.1), employing the Wald ratio method for SNP-specific estimates, which were combined using inverse-variance weighted (IVW) meta-analysis. In addition, we incorporated MR with Robust Adjusted Profile Score (MR-RAPS), a recently developed refinement designed to mitigate biases that can arise in conventional two-sample MR analyses. Additional MR methods, including MR-Egger regression, weighted median estimator, simple mode, and weighted mode, were used for validation. MR-Egger regression with Egger intercept testing was used to detect horizontal pleiotropy. Cochran’s Q test assessed heterogeneity (P < 0.05),^27,28^ and a random-effect IVW model was applied in case of significant heterogeneity. The leave-one-out method was employed to evaluate the influence of individual SNPs.

## Results

### Baseline Characteristics

A total of 475 intracranial aneurysms (IAs) from 358 patients were included, with a median age of 59 years (IQR: 52–66). The flow chart of patient selection is depicted in **Figure 1**. No significant age difference was observed between the AWE and non-AWE groups (p = 0.127). The non-AWE group had a higher proportion of females (69.2% vs. 55.6%, p = 0.005), while the AWE group had a higher smoking history (38.8% vs. 24.8%, p = 0.002). Hypertension was more prevalent in the AWE group (72.5% vs. 52.1%, p < 0.001), as was diabetes (17.5% vs. 11.4%, p = 0.091). Aneurysm characteristics differed significantly between groups: the AWE group had more internal carotid artery (ICA) aneurysms (42.5% vs. 23.8%), larger aneurysms (6.28 mm vs. 4.00 mm, p < 0.001), and a higher proportion of aneurysms ≥7 mm (43.8% vs. 11.4%, p < 0.001). Irregular aneurysm shapes (48.1% vs. 37.8%, p = 0.039) and ring-like wall enhancement (75.6% vs. 5.1%, p < 0.001) were more common in the AWE group. The AWE group also had a higher contrast ratio (CRstalk) (0.79 vs. 0.47, p < 0.001) and a higher Phases score (6.00 vs. 3.00, p < 0.001). Other detailed characteristics are detailed in **Table 1**.

**Table 1.**
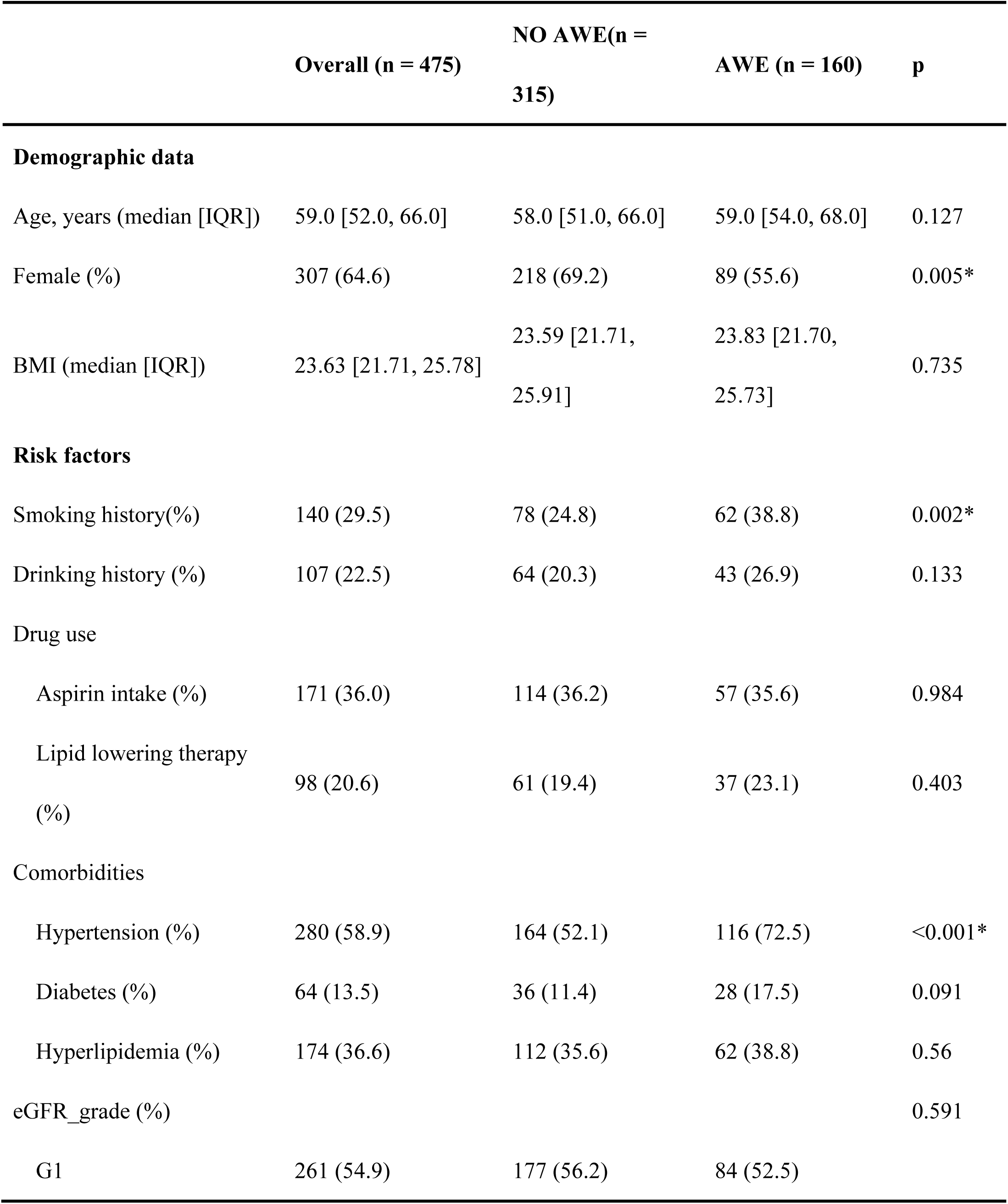
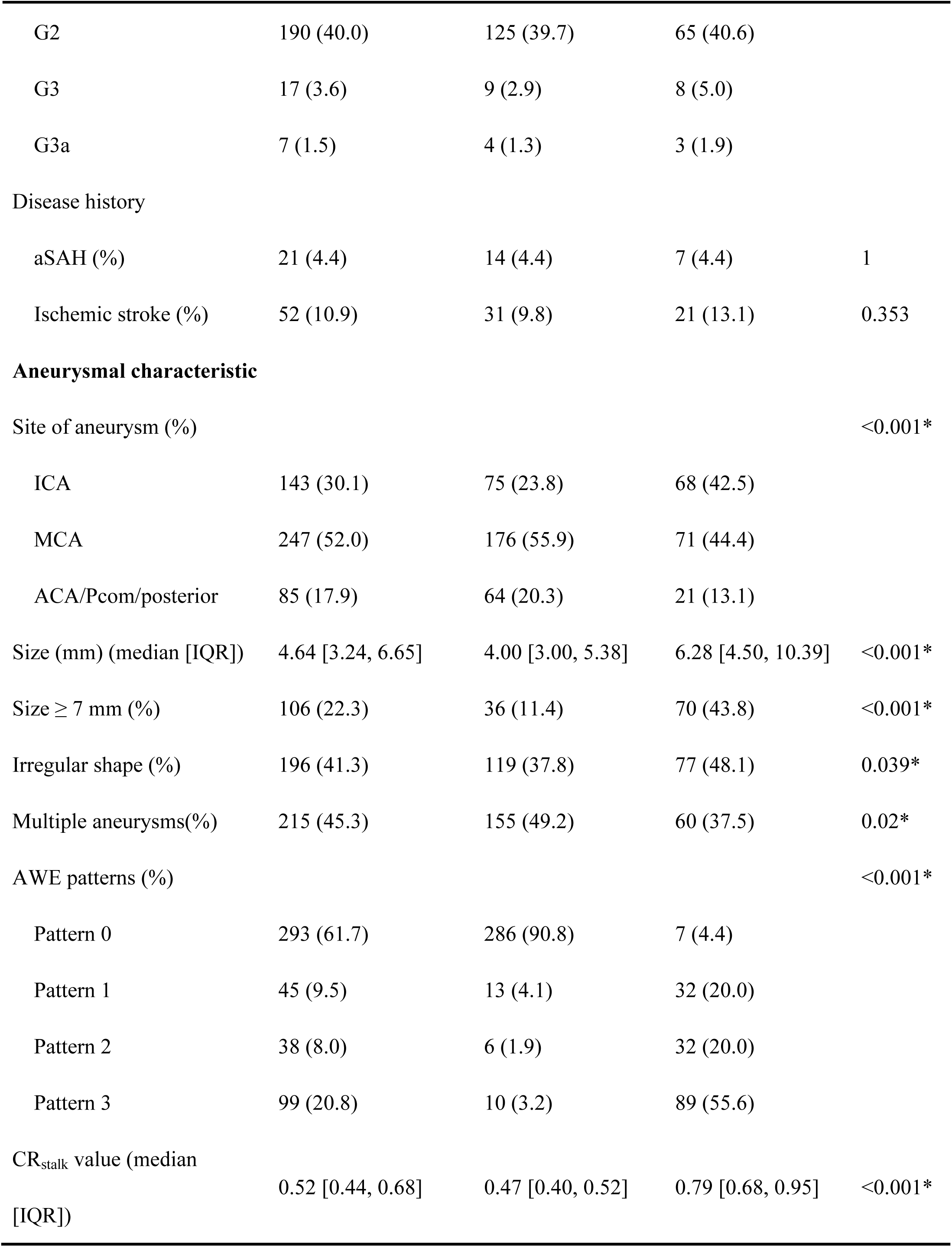
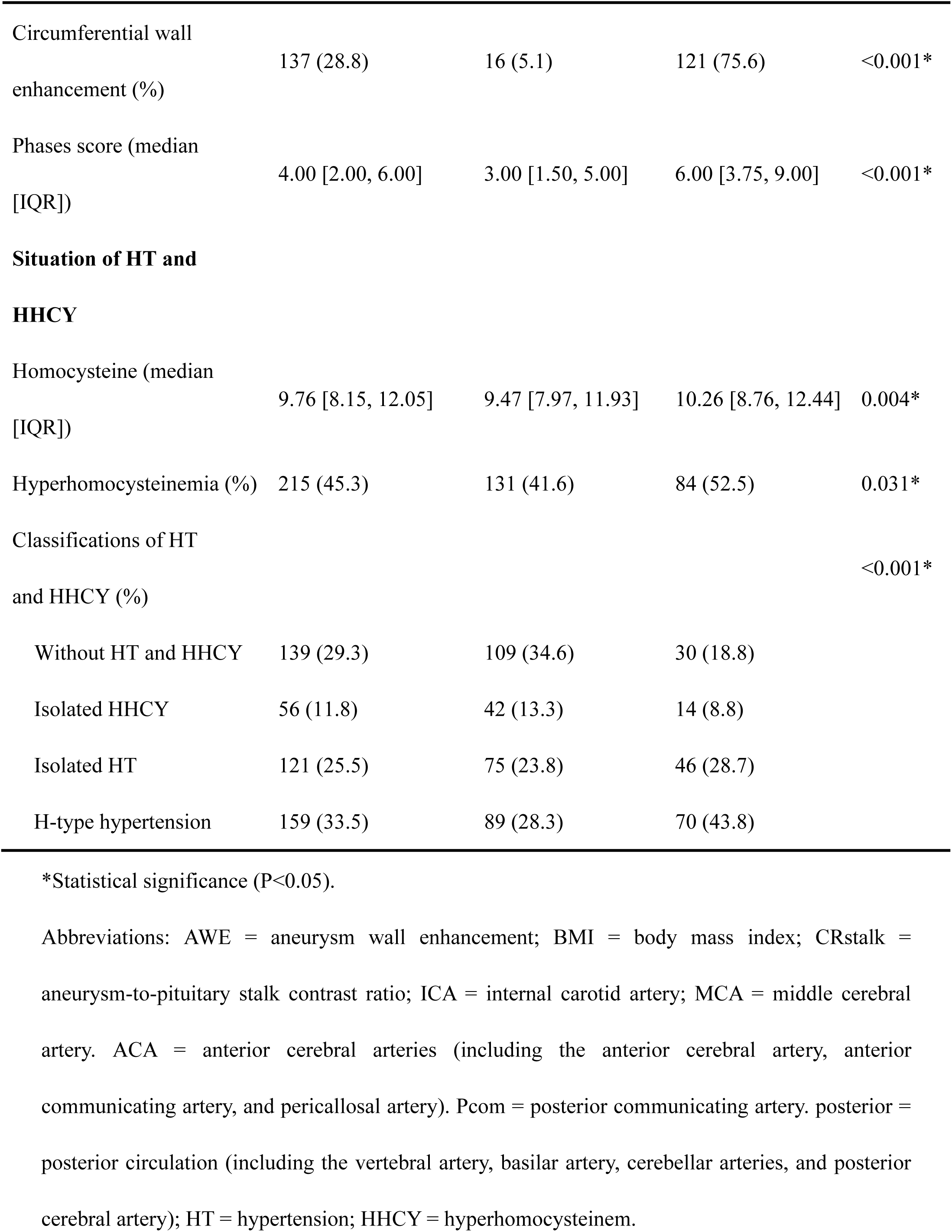
Baseline characteristics of intracranial aneurysms (IAs) patients with and without aneurysmal wall enhancement (AWE).

|  | Overall (n = 475) | NO AWE(n = 315) | AWE (n = 160) | p |
| --- | --- | --- | --- | --- |
| **Demographic data** |  |  |  |  |
| Age, years (median [IQR]) | 59.0 [52.0, 66.0] | 58.0 [51.0, 66.0] | 59.0 [54.0, 68.0] | 0.127 |
| Female (%) | 307 (64.6) | 218 (69.2) | 89 (55.6) | 0.005* |
| BMI (median [IQR]) | 23.63 [21.71, 25.78] | 23.59 [21.71, 25.91] | 23.83 [21.70, 25.73] | 0.735 |
| **Risk factors** |  |  |  |  |
| Smoking history(%) | 140 (29.5) | 78 (24.8) | 62 (38.8) | 0.002* |
| Drinking history (%) | 107 (22.5) | 64 (20.3) | 43 (26.9) | 0.133 |
| **Drug use** |  |  |  |  |
| Aspirin intake (%) | 171 (36.0) | 114 (36.2) | 57 (35.6) | 0.984 |
| Lipid lowering therapy (%) | 98 (20.6) | 61 (19.4) | 37 (23.1) | 0.403 |
| **Comorbidities** |  |  |  |  |
| Hypertension (%) | 280 (58.9) | 164 (52.1) | 116 (72.5) | <0.001* |
| Diabetes (%) | 64 (13.5) | 36 (11.4) | 28 (17.5) | 0.091 |
| Hyperlipidemia (%) | 174 (36.6) | 112 (35.6) | 62 (38.8) | 0.56 |
| eGFR_grade (%) |  |  |  | 0.591 |
| G1 | 261 (54.9) | 177 (56.2) | 84 (52.5) |  |
| G2 | 190 (40.0) | 125 (39.7) | 65 (40.6) |  |
| G3 | 17 (3.6) | 9 (2.9) | 8 (5.0) |  |
| G3a | 7 (1.5) | 4 (1.3) | 3 (1.9) |  |
| Disease history |  |  |  |  |
| aSAH (%) | 21 (4.4) | 14 (4.4) | 7 (4.4) | 1 |
| Ischemic stroke (%) | 52 (10.9) | 31 (9.8) | 21 (13.1) | 0.353 |
| **Aneurysmal characteristic** |  |  |  |  |
| Site of aneurysm (%) |  |  |  | <0.001* |
| ICA | 143 (30.1) | 75 (23.8) | 68 (42.5) |  |
| MCA | 247 (52.0) | 176 (55.9) | 71 (44.4) |  |
| ACA/Pcom/posterior | 85 (17.9) | 64 (20.3) | 21 (13.1) |  |
| Size (mm) (median [IQR]) | 4.64 [3.24, 6.65] | 4.00 [3.00, 5.38] | 6.28 [4.50, 10.39] | <0.001* |
| Size $\geq 7$ mm (%) | 106 (22.3) | 36 (11.4) | 70 (43.8) | <0.001* |
| Irregular shape (%) | 196 (41.3) | 119 (37.8) | 77 (48.1) | 0.039* |
| Multiple aneurysms(%) | 215 (45.3) | 155 (49.2) | 60 (37.5) | 0.02* |
| AWE patterns (%) |  |  |  | <0.001* |
| Pattern 0 | 293 (61.7) | 286 (90.8) | 7 (4.4) |  |
| Pattern 1 | 45 (9.5) | 13 (4.1) | 32 (20.0) |  |
| Pattern 2 | 38 (8.0) | 6 (1.9) | 32 (20.0) |  |
| Pattern 3 | 99 (20.8) | 10 (3.2) | 89 (55.6) |  |
| CR<sub>stalk</sub> value (median [IQR]) | 0.52 [0.44, 0.68] | 0.47 [0.40, 0.52] | 0.79 [0.68, 0.95] | <0.001* |
| Circumferential wall enhancement (%) | 137 (28.8) | 16 (5.1) | 121 (75.6) | <0.001* |
| Phases score (median [IQR]) | 4.00 [2.00, 6.00] | 3.00 [1.50, 5.00] | 6.00 [3.75, 9.00] | <0.001* |
| **Situation of HT and HHCY** |  |  |  |  |
| Homocysteine (median [IQR]) | 9.76 [8.15, 12.05] | 9.47 [7.97, 11.93] | 10.26 [8.76, 12.44] | 0.004* |
| Hyperhomocysteinemia (%) | 215 (45.3) | 131 (41.6) | 84 (52.5) | 0.031* |
| Classifications of HT and HHCY (%) |  |  |  | <0.001* |
| Without HT and HHCY | 139 (29.3) | 109 (34.6) | 30 (18.8) |  |
| Isolated HHCY | 56 (11.8) | 42 (13.3) | 14 (8.8) |  |
| Isolated HT | 121 (25.5) | 75 (23.8) | 46 (28.7) |  |
| H-type hypertension | 159 (33.5) | 89 (28.3) | 70 (43.8) |  |

\*Statistical significance (P<0.05).
Abbreviations: AWE = aneurysm wall enhancement; BMI = body mass index; CRstalk = aneurysm-to-pituitary stalk contrast ratio; ICA = internal carotid artery; MCA = middle cerebral artery. ACA = anterior cerebral arteries (including the anterior cerebral artery, anterior communicating artery, and pericallosal artery). Pcom = posterior communicating artery. posterior = posterior circulation (including the vertebral artery, basilar artery, cerebellar arteries, and posterior cerebral artery); HT = hypertension; HHCY = hyperhomocysteinem.

|  | Overall (n = 475) | NO AWE(n = 315) | AWE (n = 160) | p |
| --- | --- | --- | --- | --- |
| **Demographic data** |  |  |  |  |
| Age, years (median [IQR]) | 59.0 [52.0, 66.0] | 58.0 [51.0, 66.0] | 59.0 [54.0, 68.0] | 0.127 |
| Female (%) | 307 (64.6) | 218 (69.2) | 89 (55.6) | 0.005* |
| BMI (median [IQR]) | 23.63 [21.71, 25.78] | 23.59 [21.71, 25.91] | 23.83 [21.70, 25.73] | 0.735 |
| **Risk factors** |  |  |  |  |
| Smoking history(%) | 140 (29.5) | 78 (24.8) | 62 (38.8) | 0.002* |
| Drinking history (%) | 107 (22.5) | 64 (20.3) | 43 (26.9) | 0.133 |
| Drug use |  |  |  |  |
| Aspirin intake (%) | 171 (36.0) | 114 (36.2) | 57 (35.6) | 0.984 |
| Lipid lowering therapy (%) | 98 (20.6) | 61 (19.4) | 37 (23.1) | 0.403 |
| Comorbidities |  |  |  |  |
| Hypertension (%) | 280 (58.9) | 164 (52.1) | 116 (72.5) | <0.001* |
| Diabetes (%) | 64 (13.5) | 36 (11.4) | 28 (17.5) | 0.091 |
| Hyperlipidemia (%) | 174 (36.6) | 112 (35.6) | 62 (38.8) | 0.56 |
| eGFR_grade (%) |  |  |  | 0.591 |
| G1 | 261 (54.9) | 177 (56.2) | 84 (52.5) |  |
| G2 | 190 (40.0) | 125 (39.7) | 65 (40.6) |  |
| G3 | 17 (3.6) | 9 (2.9) | 8 (5.0) |  |
| G3a | 7 (1.5) | 4 (1.3) | 3 (1.9) |  |
| Disease history |  |  |  |  |
| aSAH (%) | 21 (4.4) | 14 (4.4) | 7 (4.4) | 1 |
| Ischemic stroke (%) | 52 (10.9) | 31 (9.8) | 21 (13.1) | 0.353 |
| **Aneurysmal** |  |  |  |  |

| **characteristic** |  |  |  |  |
| --- | --- | --- | --- | --- |
| Site of aneurysm (%) |  |  |  | <0.001* |
| ICA | 143 (30.1) | 75 (23.8) | 68 (42.5) |  |
| MCA | 247 (52.0) | 176 (55.9) | 71 (44.4) |  |
| ACA/Pcom/posterior | 85 (17.9) | 64 (20.3) | 21 (13.1) |  |
| Size (mm) (median [IQR]) | 4.64 [3.24, 6.65] | 4.00 [3.00, 5.38] | 6.28 [4.50, 10.39] | <0.001* |
| Size $\geq 7$ mm (%) | 106 (22.3) | 36 (11.4) | 70 (43.8) | <0.001* |
| Irregular shape (%) | 196 (41.3) | 119 (37.8) | 77 (48.1) | 0.039* |
| Multiple aneurysms(%) | 215 (45.3) | 155 (49.2) | 60 (37.5) | 0.02* |
| AWE patterns (%) |  |  |  | <0.001* |
| Pattern 0 | 293 (61.7) | 286 (90.8) | 7 (4.4) |  |
| Pattern 1 | 45 (9.5) | 13 (4.1) | 32 (20.0) |  |
| Pattern 2 | 38 (8.0) | 6 (1.9) | 32 (20.0) |  |
| Pattern 3 | 99 (20.8) | 10 (3.2) | 89 (55.6) |  |
| CR<sub>stalk</sub> value (median [IQR]) | 0.52 [0.44, 0.68] | 0.47 [0.40, 0.52] | 0.79 [0.68, 0.95] | <0.001* |
| Circumferential wall enhancement (%) | 137 (28.8) | 16 (5.1) | 121 (75.6) | <0.001* |
| Phases score (median [IQR]) | 4.00 [2.00, 6.00] | 3.00 [1.50, 5.00] | 6.00 [3.75, 9.00] | <0.001* |
| **Situation of HT and HHCY** |  |  |  |  |
| Homocysteine (median [IQR]) | 9.76 [8.15, 12.05] | 9.47 [7.97, 11.93] | 10.26 [8.76, 12.44] | 0.004* |
| Hyperhomocysteinemia (%) | 215 (45.3) | 131 (41.6) | 84 (52.5) | 0.031* |
| Classifications of HT and HHCY (%) |  |  |  | <0.001* |
| Without HT and HHCY | 139 (29.3) | 109 (34.6) | 30 (18.8) |  |

|  |  |  |  |
| --- | --- | --- | --- |
| Isolated HHCY | 56 (11.8) | 42 (13.3) | 14 (8.8) |
| Isolated HT | 121 (25.5) | 75 (23.8) | 46 (28.7) |
| H-type hypertension | 159 (33.5) | 89 (28.3) | 70 (43.8) |

Homocysteine levels were significantly higher in the AWE group (10.26 vs. 9.47 µmol/L, p = 0.004), with a higher prevalence of hyperhomocysteinemia (52.5% vs. 41.6%, p = 0.031). The AWE group also showed a higher prevalence of H-type hypertension (43.8% vs. 28.3%) compared to the non-AWE group (p < 0.001) (**Figure 3**). **Figure S1** compares the distribution of CRstalk values between patients with and without H-type hypertension (p < 0.001).

**Figure 3.**
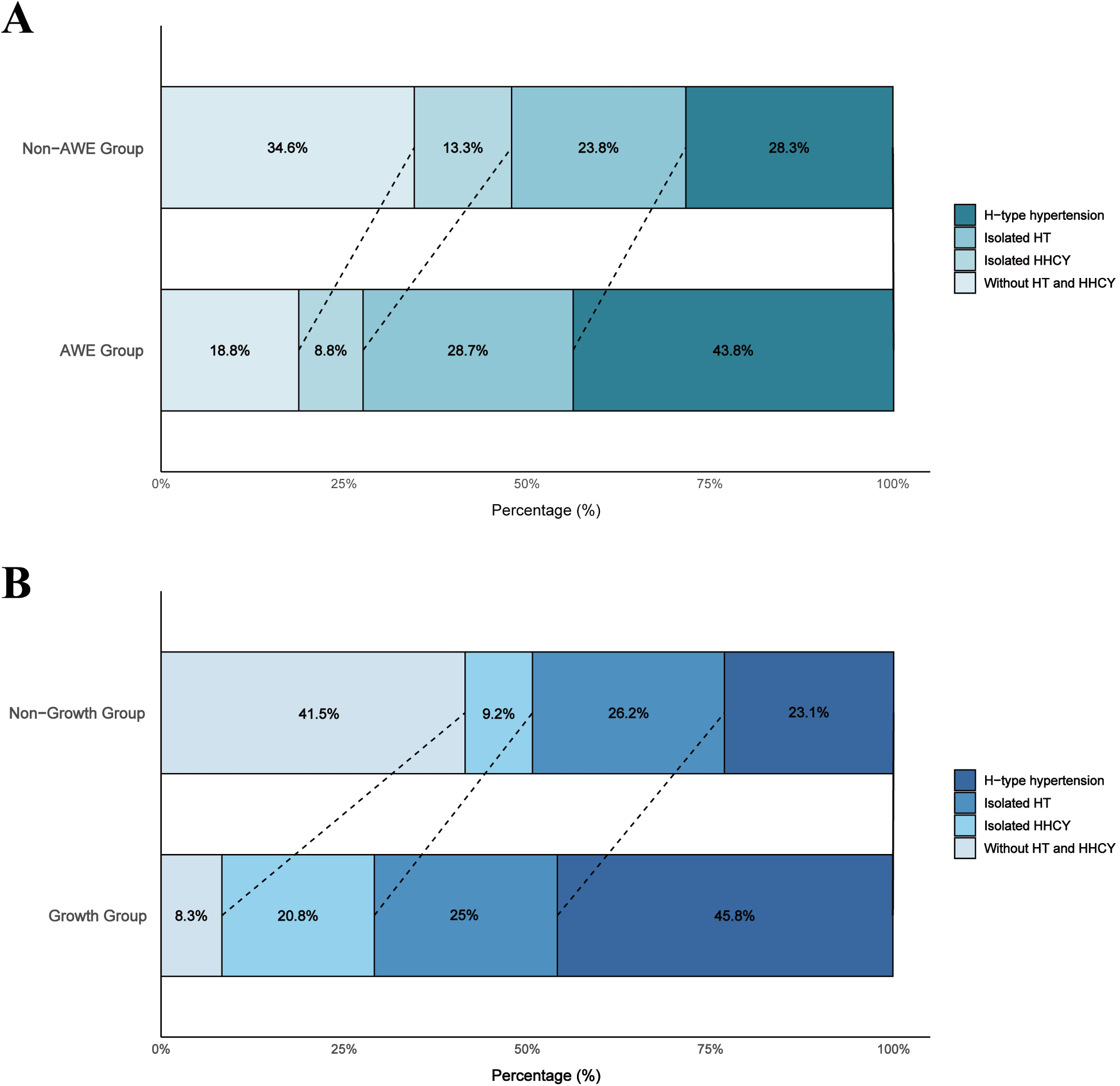
Stacked distribution of hyperhomocysteinemia and hypertension in AWE vs Non-AWE and Growth vs Non-Growth Groups. A. Comparison of hypertension and homocysteine status between the AWE and non-AWE groups. Horizontal stacked bar charts show the proportion of individuals in each of the following categories: H-type hypertension (hypertension with concurrent hyperhomocysteinemia), isolated hypertension (hypertension without hyperhomocysteinemia), isolated hyperhomocysteinemia (hyperhomocysteinemia without hypertension), and individuals without either hypertension or hyperhomocysteinemia. Dashed lines indicate shifts in category distribution between the two groups. B. Comparison of hypertension and homocysteine status between the Growth and Non-Growth groups. The same four categories are represented, with dashed lines denoting transitions in group-specific distributions. Abbreviations: AWE = aneurysm wall enhancement; HT: hypertension; HHCY: hyperhomocysteinemia.

### Single and Joint Associations of Hyperhomocysteinemia and Hypertension with AWE

In multivariable logistic regression, the association between hyperhomocysteinemia and AWE weakened with increasing adjustments. In the unadjusted model (Model 1a), hyperhomocysteinemia was significantly associated with AWE (OR = 1.552, 95% CI: 1.059–2.279), but this association diminished after adjusting for demographics (Model 1b: OR = 1.265, 95% CI: 0.826–1.937) and became non-significant in the fully adjusted model (Model 1c: OR = 1.369, 95% CI: 0.834–2.249). In contrast, hypertension remained strongly associated with AWE across all models. The unadjusted model (Model 2a) showed an OR of 2.427 (95% CI: 1.618–3.689), which remained significant after adjusting for confounders (Model 2b: OR = 2.398, 95% CI: 1.533–3.802) and in the fully adjusted model (Model 2c: OR = 2.813, 95% CI: 1.683–4.783) (**Figure 4**).

**Figure 4.**
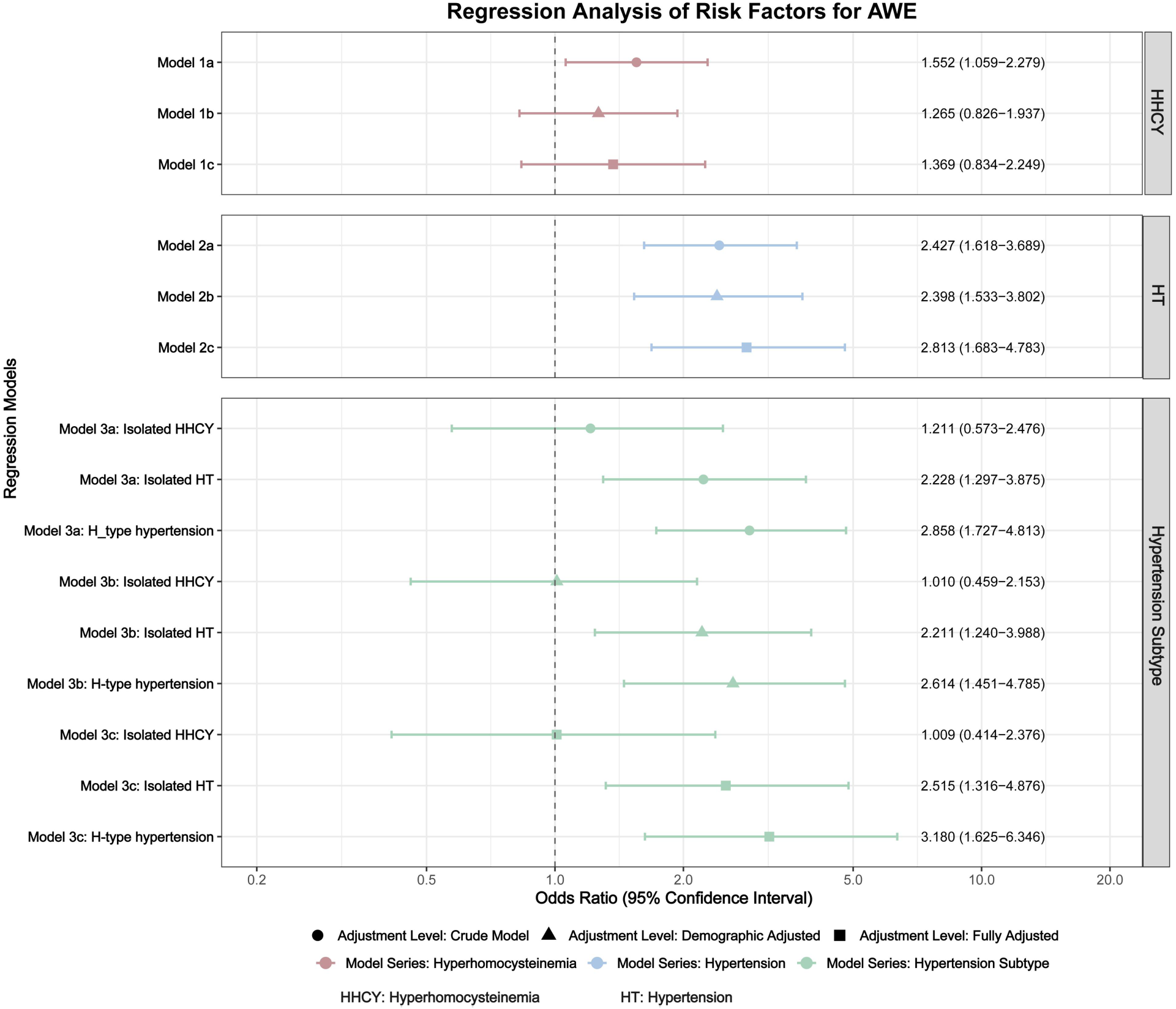
Single and joint associations of hyperhomocysteinemia and hypertension with aneurysmal wall enhancement (AWE) across regression models with incremental covariate adjustment. Odds ratios (ORs) with 95% confidence intervals (CIs) are shown for three models: hyperhomocysteinemia alone (Model 1), hypertension alone (Model 2), and their combined status (Model 3). Results are presented across three levels of adjustment: (a) unadjusted (crude), (b) adjusted for age, sex, and BMI, and (c) fully adjusted for age, sex, BMI, smoking status, aneurysm location, aneurysm size, irregular shape, and multiple aneurysms, eGFR Grade, Lipid-lowering Therapy. Symbols correspond to the adjustment level: filled circles (unadjusted), triangles (demographic-adjusted), and squares (fully adjusted). (Abbreviations: AWE = aneurysm wall enhancement; BMI = body mass index; ICA = internal carotid artery; MCA = middle cerebral artery. ACA = anterior cerebral arteries (including the anterior cerebral artery, anterior communicating artery, and pericallosal artery). Pcom = posterior communicating artery. posterior = posterior circulation (including the vertebral artery, basilar artery, cerebellar arteries, and posterior cerebral artery); HT = hypertension; HHCY = hyperhomocysteinemia.)

Joint analyses revealed that H-type hypertension had the stronger association with AWE (OR = 3.180, 95% CI: 1.625–6.346) compared to isolated hypertension (OR = 2.515, 95% CI: 1.316–4.876). Isolated hyperhomocysteinemia showed a weaker, non-significant association (OR = 1.009, 95% CI: 0.414–2.376). This pattern persisted across all models, with H-type hypertension consistently exhibiting the strongest association with AWE (**Figure 4, Table S1**).

### Subgroup and Sensitivity Analyses

The results of subgroup analyses were shown in **Table S2**. There were no significant interactions across subgroups (p for interaction>0.05). Sensitivity analyses, excluding patients with an estimated glomerular filtration rate (eGFR) < 60 (**Figure S2**), defining hyperhomocysteinemia as homocysteine ≥ 15 (**Figure S3**), or defining AWE as CRstalk ≥ 0.7 (**Figure S4**), confirmed the robustness of the findings.

### Longitudinal analysis for aneurysm growth

Among 89 aneurysms, 24 (27.0%) showed aneurysm growth during follow-up (**Table S3)**. The median follow-up interval for the prospective longitudinal cohort was 7.00 months ([IQR]: 6.00–14.00). A significant difference in follow-up duration was observed between the groups, with a median of 7.00 months (IQR: 5.00–10.00) in the non-growth group and 12.50 months (IQR: 6.75–32.25) in the growth group (p = 0.001). Significant differences in aneurysm size (4.81 mm vs. 3.55 mm, p = 0.002), aneurysm size ≥7 mm (29.2% vs. 1.5%, p < 0.001), aneurysm location (p = 0.014), and PHASES score (6.00 vs. 2.00, p < 0.001) were observed between growth and non-growth groups. Patients with growth had higher homocysteine levels (11.26 vs. 8.83 μmol/L, p = 0.026) and higher rates of hyperhomocysteinemia (66.7% vs. 32.3%, p = 0.007). H-type hypertension was more common in the growth group (45.8% vs. 23.1%, p = 0.012) (**Figure 3)**. Firth logistic regression analysis identified H-type hypertension as an independent predictor of aneurysm growth. Compared to patients without H-type hypertension, the adjusted odds ratios (ORs) were 2.775 (95% CI, 1.048–7.409; P=0.040) in Model 1, 2.693 (95% CI, 0.874–8.593; P=0.084) in Model 2, and 3.634 (95% CI, 1.058–13.514; P=0.040) in Model 3. Additionally, larger aneurysm size remained a strong independent risk factor across all models (OR, 24.630; P<0.001) (**Table S4)**. The baseline characteristics of patients with and without longitudinal follow-up imaging are compared in **Table S5**.

### Reproducibility Analyses

Interobserver agreement for the assessment of CRstalk, AWE patterns, and aneurysmal growth was assessed and is quantified in **Table S6**.

### Mendelian Randomization Analyses

The complete list of single-SNP effect estimates utilized in the TSMR analysis is provided in Supplementary **Tables S7-S9.** The analytical workflow is summarized in **Figure 1**. TSMR analysis in European-ancestry populations demonstrated that a genetically predicted one-standard-deviation increase in Hcy levels causally elevated the risk of aSAH by 38.8% (OR = 1.388, 95% CI: 1.016–1.895; p = 0.039) and increased hypertension risk by 10.2% (OR = 1.102, 95% CI: 1.010–1.201; p = 0.029), while hypertensive individuals exhibited 57.8% higher aSAH risk compared with normotensive controls (OR = 1.578, 95% CI: 1.361–1.829; p < 0.001). Moreover, under the MR-RAPS approach these causal relationships remained significant or borderline-significant, indicating that our findings are minimally influenced by weak instruments and pleiotropy. The forest plot summarizing the key MR results is presented in **Figure 5**. Detailed results of the three MR analyses are provided in **Table S10**. Cochran’s Q test indicated no significant heterogeneity among the IVs used in any MR analysis (p > 0.05). Similarly, the MR-Egger intercept test revealed no evidence of horizontal pleiotropy (p > 0.05). Critically, the MR-PRESSO-corrected dataset showed no distortion from outliers, with no significant SNPs excluded during analysis, further confirming the robustness of our findings. The results of the sensitivity analyses are presented in **Table S11**. Visualization results from the scatter plots, funnel plots, and leave-one-out sensitivity analyses are presented in **Figure 6**. However, the two-step MR mediation analysis did not support a causal pathway whereby plasma Hcy influences aSAH risk through hypertension mediation (P = 0.202), with detailed mediation results provided in Supplementary **Table S12**.

**Figure 5.**
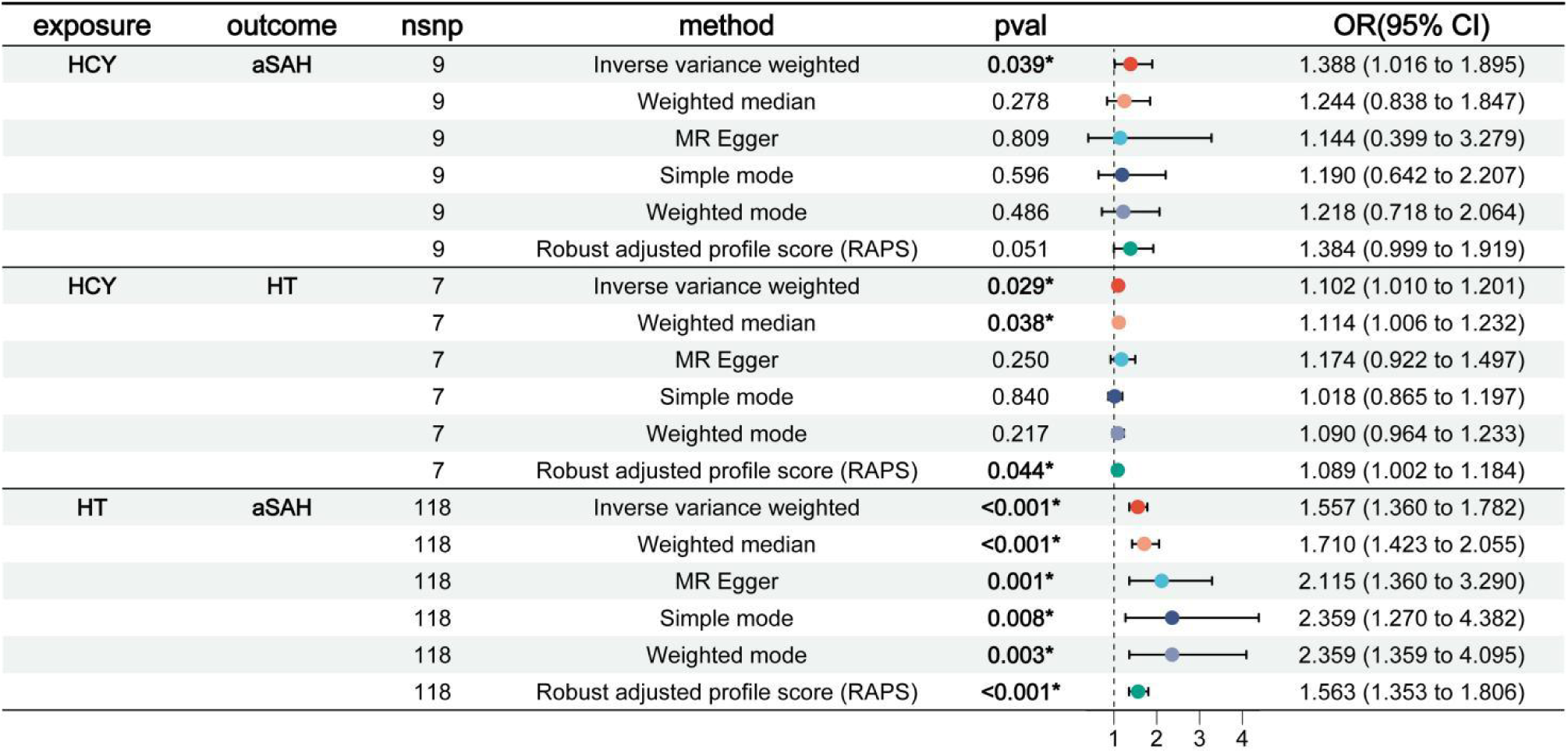
MR Analysis confirmed the causal association between Hcy, hypertension, and aSAH. Estimation of the causal association between Hcy, hypertension, and aSAH using different MR analysis methods. OR indicates odds ratio; and SNP, single-nucleotide polymorphism.

**Figure 6.**
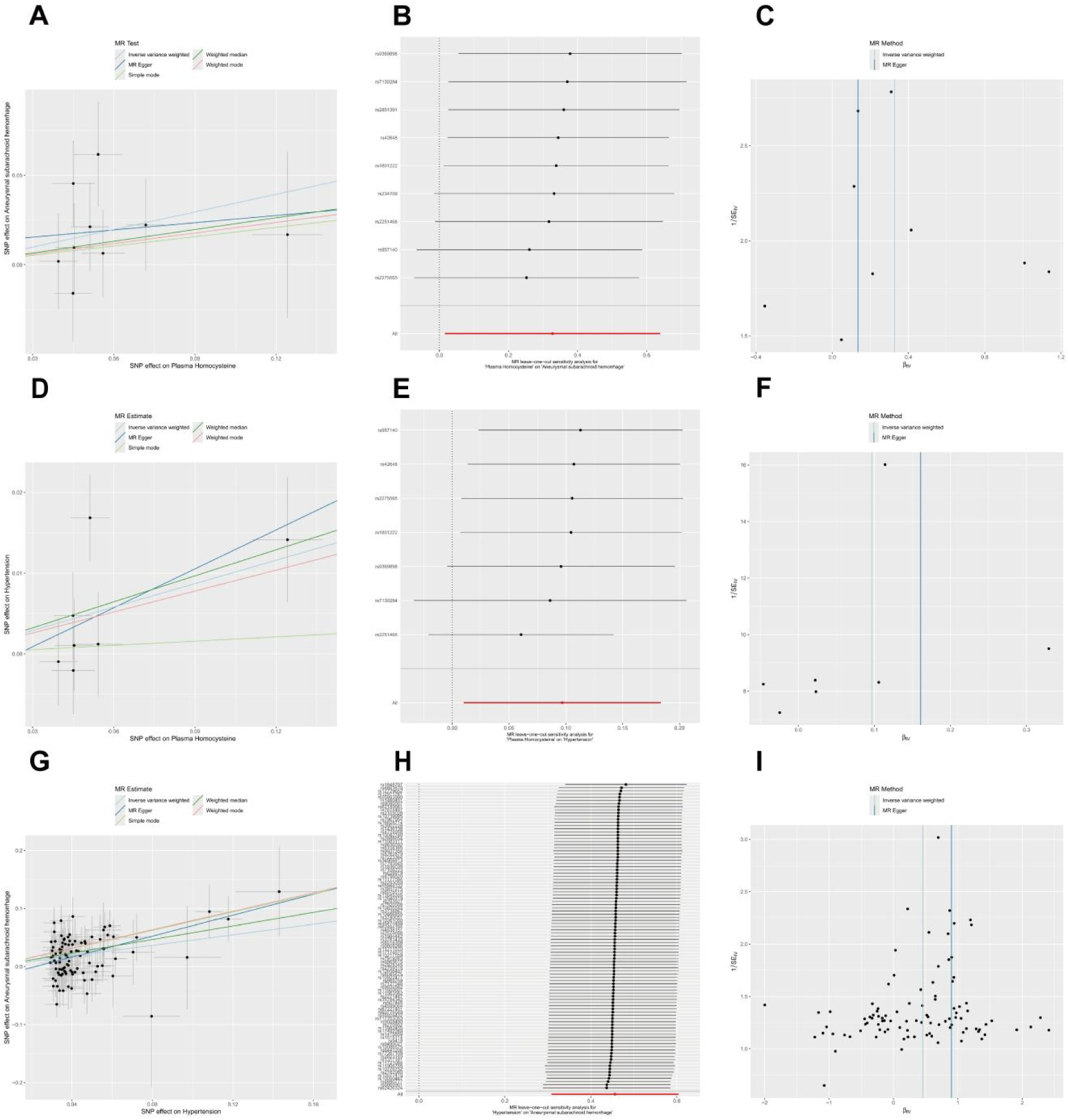
Scatter plots: A, D and G; leave-one-out sensitivity analyses plots: B, E and H; funnel plots: C, F and I. A. The scatter plot presents the results of the association analysis between plasma Hcy levels and aSAH using five MR analytical methods. B. The leave-one-out sensitivity analysis demonstrated that the effect of plasma Hcy levels on aSAH was robust to the exclusion of any individual SNP. C. Absence of outliers is demonstrated in the funnel plot assessing the effect of plasma Hcy levels on aSAH. D. The scatter plot presents the results of the association analysis between plasma Hcy levels and hypertension using five MR analytical methods. E. The leave-one-out sensitivity analysis demonstrated that the effect of plasma Hcy levels on hypertension was robust to the exclusion of any individual SNP. F. Absence of outliers is demonstrated in the funnel plot assessing the effect of plasma Hcy levels on hypertension. G. The scatter plot presents the results of the association analysis between hypertension and aSAH using five MR analytical methods. H. Leave-one-out sensitivity analysis confirmed that the causal effect of hypertension on aSAH remained robust upon exclusion of any individual SNP. I. Absence of outliers is demonstrated in the funnel plot assessing the effect of hypertension on aSAH.

### Proteomics Data Analysis

In the deCODE database, we observed nominal associations between homocysteine (Hcy) levels and the expression of 127 circulating proteins, as detailed in **Table S13**. Similarly, in the UK Biobank Pharma Proteomics Project (UKB-PPP), nominal associations were identified between Hcy and 90 plasma proteins, with results provided in **Table S14**. A secondary analysis conducted using the same platform in the Finnish cohort revealed 51 plasma proteins exhibiting nominal significance; further details are available in Supplementary **Table S15**. However, when applying the false discovery rate (FDR) method for multiple test correction across all datasets, no proteins demonstrated significant associations (FDR > 0.05). To identify biologically relevant pathways, we integrated results from three proteomic databases and performed a comprehensive GO/KEGG enrichment analysis. This analysis focused on the top 50 proteins with the smallest P-values (P < 0.05), a conventional threshold for such analyses when few proteins meet stricter FDR criteria. All pathways reported were significant at FDR < 0.05 (or q-value < 0.05). Key genes (e.g., IL17RA, TSLP, SLAMF1, PRTN3) were recurrently enriched across multiple pathways, pointing to four core biological themes: inflammatory-immune dysregulation, immune-cell recruitment, extracellular-matrix remodelling, and signalling-pathway activation. These themes constitute the central mechanisms through which Hcy likely mediates aSAH risk. Full results are available in **Table S16, Figure S5 and Figure S6.**

## Discussion

We provide the first comprehensive evidence elucidating the intricate interplay between hypertension and hyperhomocysteinemia in intracranial aneurysm (IA) instability. By integrating multi-modal data—including observational metrics, high-resolution vessel wall MRI (HRVWI), Mendelian randomization (MR), and proteomic profiling—we demonstrate that H-type hypertension, defined as the co-occurrence of hypertension and elevated Hcy, is significantly associated with aneurysm growth and wall enhancement (AWE), an important indicator of wall inflammation and rupture risk. MR analysis further supports a causal role of Hcy in promoting both hypertension and IA rupture. Our findings offer critical implications for risk stratification and clinical management of patients with unruptured intracranial aneurysms, while enhancing the mechanistic understanding of aneurysm pathogenesis.

Hypertension has long been recognized as a key independent risk factor for IA rupture.^3–5^ Consistent with previous evidence, our study confirmed this association across multiple multivariable models. Importantly, our data also highlighted the augmented risk associated with H-type hypertension, which exhibited a stronger association with AWE compared to isolated hypertension. Specifically, joint effect analysis demonstrated that the odds ratio (OR) for AWE in patients with H-type hypertension was 3.075, compared to 2.426 in those with hypertension alone. This suggests a potential synergistic effect, whereby the coexistence of elevated blood pressure and Hcy levels may amplify vascular wall stress and inflammatory injury,^9^ contributing to aneurysm destabilization. Consistent results were observed in our longitudinal cohort: patients with H-type hypertension had a higher rate of aneurysm growth (45.8% vs. 23.1%). In multivariable logistic regression analysis, H-type hypertension remained independently associated with aneurysm growth (OR, 21.743; P = 0.020). These findings underscore the importance of considering H-type hypertension as a distinct risk factor in the management of patients with intracranial aneurysms, emphasizing the need for targeted strategies aimed at both blood pressure and Hcy levels to reduce aneurysmal rupture risk.

Previous studies have suggested that elevated homocysteine (Hcy) levels are associated with an increased risk of intracranial aneurysm (IA) rupture,^29–31^ with higher serum homocysteine concentrations correlating with greater aneurysmal wall enhancement (AWE) in fusiform aneurysms.^18^ In our univariate analysis, elevated homocysteine (Hcy) was significantly associated with aneurysmal wall enhancement (AWE), a marker indicative of aneurysmal inflammation and increased rupture risk. However, this association weakened in multivariable models, likely due to the confounding influence of hypertension. Prior studies have demonstrated a positive correlation between plasma Hcy levels and the incidence of hypertension and elevated blood pressure, suggesting that hypertension may mediate or confound the relationship between homocysteine and aneurysmal instability.^32,33^

To further explore this relationship, we performed Mendelian randomization (MR) analysis, which provided deeper insights into the causal links between Hcy, hypertension, and aneurysmal rupture. Although the mediation effect was not statistically significant, our Mendelian randomization analysis demonstrated that genetically determined homocysteine (Hcy) levels were positively associated with both hypertension and aneurysmal rupture. Moreover, hypertension itself was significantly linked to an increased risk of rupture. These findings suggest that Hcy may contribute to aneurysmal rupture through both direct and indirect pathways. Indirectly, elevated total Hcy levels may destabilize the aneurysmal wall, at least in part, by exacerbating hypertension. Elevated homocysteine (Hcy) levels induce oxidative stress by increasing reactive oxygen species (ROS), which damages endothelial cells and reduces nitric oxide (NO) bioavailability, leading to vasoconstriction.^34,35^ Hcy also stimulates vascular smooth muscle cell (VSMC) proliferation and migration, causing vascular wall thickening and increased peripheral resistance.^36^ Additionally, Hcy has pro-inflammatory effects, activating inflammatory pathways that impair endothelial function and promote atherosclerosis, thereby worsening hypertension.^37^ Hcy may also enhance angiotensin II production or its effects, further elevating blood pressure.^38,39^ Another key mechanism is genomic DNA hypomethylation. Elevated Hcy reduces DNA methylation, leading to the demethylation and upregulation of hypertension-related genes, contributing to elevated blood pressure.^40–42^

Furthermore, hypertension heightens the risk of intracranial aneurysm rupture by increasing hemodynamic stress and driving vascular wall remodeling.^3,43,44^ Beyond directly increasing hemodynamic stresses, hypertension activates both systemic and local renin-angiotensin systems (RAS),^45^ which initiate vascular inflammation, endothelial dysfunction, and NF-κB-mediated signaling.^46^ These factors promote smooth muscle cell migration and proliferation, ultimately destabilizing the aneurysmal wall.^47,48^ On the other hand, directly, Hcy may exert harmful effects on the vascular endothelium through mechanisms such as oxidative stress and inflammation, even in the absence of hypertension.^6,49,50^ These pathways can lead to endothelial dysfunction, matrix degradation, and vascular wall weakening, all of which increase the risk of aneurysm rupture.^51^

Notably, in the context of H-type hypertension, these pathogenic pathways may not simply add up but rather act synergistically to amplify vascular injury. Hypertension increases hemodynamic wall stress and activates pro-inflammatory and remodeling pathways (e.g., RAS and NF-κB signaling).^52^ Concurrent hyperhomocysteinemia further lowers the threshold for oxidative stress, exacerbates endothelial dysfunction, and promotes inflammatory cascades, thereby magnifying vascular damage.^53^ This mutual reinforcement establishes a pathological positive feedback loop, potentially accelerating aneurysmal wall inflammation and adverse remodeling, and disproportionately increasing rupture risk beyond the effect of either factor alone.^54,55^ This framework of synergistic interaction is strongly supported by our integrative proteomic enrichment analyses. By integrating results from three major proteomic databases, we identified molecular mechanisms linking Hcy to aneurysmal rupture, converging on four core biological themes: inflammation-immune dysregulation, immune cell recruitment, extracellular matrix remodeling, and aberrant pathway activation.^56–59^ Key genes (e.g., IL17RA, TSLP) were cross-enriched in multiple pathways, including leukocyte migration, cytokine-cytokine receptor interaction (hsa04060), and JAK-STAT signaling (hsa04630), providing direct molecular evidence for Hcy-driven persistent vascular wall inflammation and matrix degradation.

The clinical implications of our findings are noteworthy. H-type hypertension may represent a high-risk vascular phenotype in patients with unruptured IAs, warranting early identification and aggressive risk factor modification. Management strategies should include optimal blood pressure control and Hcy-lowering interventions, such as folic acid and B-vitamin supplementation. This approach is supported by evidence from the China Stroke Primary Prevention Trial (CSPPT), which demonstrated a reduction in stroke incidence among individuals with H-type hypertension treated with folate.^60–62^ However, while these findings suggest potential benefits, the hypothesis regarding the impact of folic acid supplementation on aneurysm rupture risk is derived indirectly from observational and genetic associations. The specific effects of folic acid on aneurysm rupture risk remain to be directly tested, and require validation in future randomized controlled trials specifically designed with intracranial aneurysm growth or rupture as the primary endpoint. Eligible patients with unruptured intracranial aneurysms and H-type hypertension would be randomized to receive either standard management (guided by current hypertension guidelines) or intensive dual management (combining stringent blood pressure control and homocysteine-lowering therapy, e.g., with folic acid, vitamin B12, and B6). The primary endpoint would be a composite of aneurysm rupture or progressive growth (≥1 mm) on scheduled follow-up high-resolution vessel wall MRI, performed at baseline, 6 months, and 12 months. Such a trial would provide the highest level of evidence needed to evaluate this potential preventive strategy. Furthermore, for patients with IAs and H-type hypertension, intensified imaging surveillance using high-resolution vessel wall MRI may be warranted to monitor inflammatory changes and aneurysmal growth. Moreover, future longitudinal studies incorporating serial biomarker measurements could better elucidate the dynamic relationship between Hcy levels, blood pressure control, and aneurysm progression.

Although nominal associations between homocysteine (Hcy) and circulating protein expressions were observed in several large databases, none of these associations remained significant after applying false discovery rate (FDR) correction. This suggests that the impact of Hcy on aneurysmal subarachnoid hemorrhage (aSAH) may not be mediated through these proteins, but rather through other molecular mechanisms not captured in the current study. While these preliminary findings provide clues for potential relationships between Hcy and protein expression, the nominal associations observed without multiple testing correction are unlikely to have biological significance. Therefore, future research should incorporate larger sample sizes and more refined analytical approaches to further validate these associations and explore the underlying mechanisms of Hcy in cardiovascular and neurological diseases.

## Limitations

Several limitations of this study should be acknowledged. First, the longitudinal analysis was constrained by a modest sample size and non-random, clinically determined inclusion of conservatively managed patients, which may limit statistical power and introduce selection bias. The differential follow-up duration between the growth and non-growth groups, while reflective of real-world surveillance, could further contribute to surveillance bias. Second, the lack of detailed hypertension phenotyping—such as hypertension gradesor control status—restricted further mediation analysis of its potential role in aneurysmal instability. Third, the homocysteine threshold (≥10 μmol/L), although commonly used in Asian populationsand supported by sensitivity analyses, may not be generalizable to other ethnic groups due to inter-ethnic differences in genetics, dietary habits, and folate metabolism. Fourth, in the Mendelian randomization (MR) analysis, the limited number of genetic instruments for homocysteine reduced the sensitivity to detect horizontal pleiotropy. Moreover, while MR supports a potential causal role, it does not delineate the specific underlying biological pathways. Finally, several methodological disparities may also affect interpretation and clinical translation. These include ethnic differences between the observational cohort (primarily Asian) and genetic datasets (predominantly European), the use of aneurysmal subarachnoid hemorrhage (aSAH) as the primary MR outcome, and the reliance on surrogate markers—such as aneurysm wall enhancement and growth—rather than rupture in the observational cohort. This design limits the direct inference of our findings to the risk of aneurysm rupture, which warrants confirmation in future studies with clinical rupture as a primary outcome. Although our mechanistic interpretation of the proteomic findings is supported by existing literature, we urge readers to treat it cautiously; definitive experimental and large-scale observational evidence is still required. Given current constraints, our study offers the best-available explanation for these associations.

Looking ahead, the integration of multimodal data—including imaging, genetic, proteomic, and clinical metrics—through artificial intelligence (AI) frameworks holds promise for refining risk stratification in unruptured intracranial aneurysms and could ultimately support personalized monitoring schedules and clinical decision-support systems.^63^

## Conclusions

This study provides the first comprehensive assessment of H-type hypertension’s impact on aneurysmal instability and growth, confirming a causal relationship. These findings may have clinical implications for risk stratification and management of UIA patients, offering new insights into the underlying disease mechanisms.

## Contribution statement

All authors contributed to the paper, satisfying the ICMJE guidelines for authorship. Jiwan Huang, Hao Yuan and Ruiyan Liu: designed, conceptualized the study and drafted the manuscript; Xueyan Deng, Chi Huang, Mengshi Huang and Zhuohua Wen: analyzed and interpreted the data; Caihong Li, Yuxin Li, Lele Dai, Wenxin Chen, Bohan Cai, Runze Ge, Can Li, Jiancheng Lin, Yi Tu, Yiming Bi, Fa Jin and Shixing Su: collected the data; Xiru Zhang, Shuyin Liang, Canzhao Liu, Xin Zhang and Xifeng Li: critically revised the study outcomes; Zhibo Wen, Chuanzhi Duan and Xin Feng: funding, study supervision, and critical revision of the manuscript.

## Ethics approval

This study involves human participants and the protocol was approved by local Ethical Committees of Zhujiang Hospital of Southern Medical University (2023-KY-038-03). Participants gave informed consent to participate in the study before taking part.

## Patient consent for publication

Not applicable. The authors have no personal, financial, or institutional interest in any of the drugs, materials, or devices described in this article.

## Provenance and peer review

Not commissioned; externally peer reviewed.

## Data availability statement

Data are available upon reasonable request. The data supporting the findings of this study are available from the corresponding author upon reasonable request.

## What Is New?

Hypertension and hyperhomocysteinemia are known to contribute to aneurysm rupture, yet the impact of H-type hypertension (coexisting hypertension and elevated homocysteine) on aneurysmal instability is unclear. Using a large cohort with high-resolution vessel wall MRI, we identify H-type hypertension as an independent risk factor for aneurysmal wall enhancement and growth. Mendelian randomization confirms a causal relationship between homocysteine, hypertension, and rupture risk. Proteomic analysis reveals potential biological mechanisms underlying these associations.

## What Is Relevant?

Highlights the synergistic effect of hypertension and hyperhomocysteinemia on aneurysmal wall instability. Provides evidence linking H-type hypertension with higher risks of aneurysm growth and rupture.

## Clinical/Pathophysiological Implications

Managing elevated homocysteine in hypertensive patients may reduce aneurysm rupture risk by stabilizing the aneurysm wall, supporting updated clinical guidelines for homocysteine management in high-risk groups.

